# Real-world study of Cerviron^®^ vaginal ovules in the treatment of cervical lesions of various etiologies

**DOI:** 10.1101/2023.05.09.23289651

**Authors:** Izabella Petre, Daniela Teodora Sirbu, Ramona Petrita, Andreea-Denisa Toma, Ema Peta, Florentina Dimcevici-Poesina

**Affiliations:** Discipline of Obstetrics and Gynecology XII, ‘Victor Babes’ University of Medicine and Pharmacy, 300041 Timisoara; University Clinic of Obstetrics and Gynecology, Emergency Clinic Hospital ‘Pius Brinzeu’, 300226 Timisoara; Dr Sirbu Daniela Private Practice, 300269 Timisoara; Biometrics Unit, MDX Research, 300633 Timisoara; Pharmacy Unit of Municipal Hospital ‘Dr Karl Diel’ Jimbolia, 305400 Jimbolia; Quality Assurance, Perfect Care Distribution SRL, Bucharest, Romania

**Keywords:** medical device, cervical lesions, cervical mucosa, vaginal pH, vaginal pain, vaginal bleeding

## Abstract

Cervical lesions can be caused by pathogens, hormonal changes or by cervical injury. The recommended treatment in all cases is excision. Local re-epithelialization therapy should be initiated preoperatively and postoperatively. The present study assessed the post-market performance and tolerability of Cerviron^®^ ovules in the treatment and management of cervical lesions postoperatively. The study population included 345 participants aged 20-70 years with either a cervical lesion under treatment or with recent surgical removal of a cervical lesion. The degree of re-epithelialization of the cervical mucosa was improved in 73.17% of the patients evaluated during routine colposcopy exams and 92.73% of patients recorded no bleeding. When adding Cerviron^®^ either as monotherapy or in association with other antimicrobials in postoperative care of the cervical ectropion, improved postoperative outcomes such as reduced post-interventional bleeding and a superior quality of healing were observed. The study and its details are registered in www.clinicaltrials.gov under ID NCT05668806.

## Introduction

A cervical erosion (or cervical ectropion) can occur for numerous reasons among women of childbearing age (1). It is considered a benign condition caused by pathogens or induced by increased exposure of the cervical epithelium to estrogen. Moreover, cervical erosions can be of traumatic, mechanical etiology, such as intrauterine devices or foreign bodies introduced into the vagina (2). Its prevalence ranges from 17 to 50% (3,4). The presence of ectropion is detected especially after the menarche, during pregnancy or with use of the combined oral contraceptive pill and is very rare in postmenopausal women (5).

Epithelialization of the vaginal mucus and cervix is crucial in the management of cervical erosions. Acute inflammation of the cervix as a result from direct infection or trauma translates into several symptoms, such as white to yellow vaginal discharge (predominant symptom caused by the mucus-secreting glandular epithelium), post-coital or intermenstrual bleeding, dysuria, pelvic pain, vulvovaginal irritation and dyspareunia (6). Symptomatic women should be screened for infective agents (7). Cervical ectropion has been associated with both the combined oral contraceptive pill and intrauterine contraceptive devices as highlighted by the study conducted by Wright *et al* (4). Concurrently, a number of studies have highlighted that the use of combined oral contraceptives is highly associated with the development of cervical ectopy, edema and erythema of the ectopic zone (7-9).

Pathogens such as streptococci, staphylococci, or enterococci can be promoters of acute inflammation of the cervix and cervical ectropion (10). Cervicitis (inflammation of the cervix) is often asymptomatic and can cause complications of the upper genital tract with ectopia and cervical infection by *Chlamidia trachomatis, Neisseria gonorrhoeae*, herpes simplex virus, and cytomegalovirus (11). Some factors involved in the pathogenesis of cervical ectopy involve the action of estrogen (12). Estrogens influence immune and inflammatory processes, by regulating chemokines and chemokine receptors. In a previous study by Straub, the complex processes of inflammation related to estrogen signaling toward immune cell trafficking were studied (13). The T helper 17 cells producing IL-17 are the main T cells responsible for chronic inflammation. The cervical epithelium is highly responsive to estrogen production. Mechanistically, it is considered that estrogens induce apoptosis in cervical cells and also increase gene expression of human papillomavirus-16 and -18, the two genotypes frequently associated with cervical cancer (14). Exposure to high levels of estrogen is also linked to an increased risk of breast cancer (15). Estrogen influences immune and inflammatory processes by modulating the production of pro-inflammatory cytokines, chemokines and other immune mediators. Specifically, estrogen can activate pathways that lead to the production of anti-inflammatory cytokines, such as interleukin-4 (IL-4), and chemokines, such as interferon-γ (IFN-γ) (16). Furthermore, estrogen can inhibit the production of pro-inflammatory mediators such as tumor necrosis factor (TNF) and IL-6. Estrogen can also regulate the expression of chemokine receptors on immune cells, which in turn helps regulate the migration of immune cells to sites of inflammation (17). For example, estrogen can downregulate the expression of the CCR5 receptor, which is involved in the migration of T cells to sites of inflammation. Estrogen can also upregulate the expression of the CXCR4 receptor, which is involved in the migration of macrophages and neutrophils to sites of inflammation (18).

The presence of endocervical columnar epithelium on the ectocervix favors an increased exposure to infections due to low cell-mediated immunity. In these areas, the subpopulation of T lymphocytes, namely, T helper cells, CD8 cells, and CD1 lymphocytes are reduced in number (19). Therefore, the columnar epithelium cells are more susceptible to infections such as *Chlamydia trachomatis* or *Neisseria gonorrhoeae* (20). Estrogens are capable of markedly altering the responses of host cells to microbes. In adolescence, pregnancy, during hormonal contraception, or during the years of menstruation (mostly in the ovulatory phase), the probability of developing cervical ectopy is very high, and sometimes goes undetected (5). Furthermore, cervical ectopy implies further risks of acquiring sexually-transmitted diseases (gonorrhea, chlamydia and human papilloma virus). In the study conducted by Sanchez *et al* a causal relationship was found between cervical erosion and bacterial vaginosis that alter the mucosal barrier and decrease defense mechanisms of the cervix and vagina (21).

An ongoing debate remains of whether ectopy requires a specific treatment. The association between squamous metaplasia and induction of squamous cell carcinoma of the cervix is well known. Moreover, the dysplastic cells are more susceptible to carcinogens (22). Notwithstanding this, according to a recent study conducted by Kleppa *et al*, ectopy may be a biological risk factor for chlamydia infection and for human immunodeficiency virus (HIV) in adolescents and in young women (7).

Currently, cryotherapy (cryosurgery) of the cervix is the standard treatment for symptomatic, benign cervical ectropion (23). Cryosurgery improves the cervical mucus characteristics and therefore it is recommended in patients with hostile cervical mucus and ectropion (24). Prior to and after surgery, adjuvant treatments should be promoted with the support of local, re-epithelizing treatments (25). For example, Belfiore *et al* reported the effectiveness of a topical treatment for cervical ectropion with 5 mg of deoxyribonucleic acid (26).

Cerviron has a substance content that provides beneficial properties in non-infectious 100 vulvovaginitis and cervical erosion. The ovule melts in the vaginal mucosa forming a cream that 101 ensures dispersion of the substances contained and acts as a protective barrier with astringent 102 effect, favoring the reepithelization of damaged tissue and the restoration of the initial 103 colpoecosystem without affecting the Doderlein bacilli. The main mechanism of action of the 104 medical device is the dispersion of the substances in the vagina and the formation of a protective 105 barrier that accelerates the natural healing process of the damaged epithelium. Performance and safety data on Cerviron^®^ vaginal ovules have been reported from multiple sources. Three clinical investigations on Cerviron were reported (NCT04735705, NCT04735718 and NCT05652959 available at https://clinicaltrials.gov/ct2/show/NCT04735705; https://clinicaltrials.gov/ct2/show/NCT04735718; and https://clinicaltrials.gov/ct2/show/NCT05652959, respectively) and published articles on Cerviron^®^ include the characterization of its utility in the management of cervical uterine fibroids and on vaginal atrophy after surgical treatment and adjuvant radiation therapy for cervical cancer (27,28). However, previous safety data was limited and included only a restricted, homogenous population, including 50 participants in study NCT04735705 and only 27 participants in study NCT04735718.

Cerviron^®^ is a medical device marketed by Perfect Care Distribution in the following countries: Albania, Latvia, Lithuania, Estonia, Kosovo, Montenegro, Romania, Kuwait, and the United Arab Emirates. With a complex composition consisting of three topical pharmaceutical products including hexylresorcinol, collagen and bismuth subgallate, and four phytotherapeutic extracts including *Calendula officinalis, Hydrastis canadensis, Thymus vulgaris* extract and *Curcuma longa*, it is intended as adjuvant treatment for the management of cervical lesions and vulvovaginitis.

## Materials and methods

### Study objectives

The present study was designed as part of the medical device post-marketing clinical follow-up, involving routine care from a variety of clinical practices. The study includes an open-label, multicentric, non-randomized, single-arm, real-world evidence study design. The data were collected between the 20th of May 2021 and 31st of July 2021.

Real-world evidence studies are post-marketing studies bringing valuable information related to the medical devices’ safety and performance profiling and a broader understanding of the practice pattern and the clinical outcomes. The rationale of the study was aimed at capturing safety data in a broader, more heterogenous population. Cerviron^®^ vaginal ovules have been used with success in the treatment of acute and chronic vulvovaginitis of mechanical etiology, and in cervical lesions of mechanical origin, but with a limited number of study participants (NCT04735705 and NCT04735718). Real-world evidence studies and clinical trials are complementary. The present study is considered a real-world evidence study as it reflects actual clinical aspects with data collected in the context of routine delivery of care, as opposed to data collected within a clinical trial, where study design controls variability in ways that are not representative of real-world care and outcomes.

The primary objective was to evaluate the tolerability of Cerviron^®^ ovules in the treatment and management of cervical erosions of various etiologies. The secondary objective of this study was the assessment of performance of the medical device by clinical exam and patients’ degree of satisfaction related to the use of the medical device.

### Study population

The target population included women aged 20 to 70 years with symptomology associated with cervical ectropion of various etiology such as cervix trauma, postpartum injuries, vaginal infections and cervicitis. A total of 345 women were evaluated. A number of patients were treated with Cerviron^®^ ovules as monotherapy (n=210) and other patients were prescribed Cerviron^®^ ovules as an adjuvant in therapeutic schemas containing antibiotics, antivirals and/or anti-inflammatory drugs (n=135). Subjects with a previous history of any malignancy, including subjects with vulvar, vaginal, or cervical cancer or with undiagnosed abnormal genital bleeding were excluded.

The study involved 30 Romanian specialist physicians as investigators from 16 institutions each treating between 6 and 22 patients. The participating clinical practices and their locations are listed as Table 1. Upon study entry and at 1, 2 and 3 months after the initial visit, participants were interviewed and received visual cervical examinations by colposcopy. At each visit, participants received a standardized pelvic exam with placement of a speculum and visualization of the cervix.

**Table 1.** Clinical practices and their locations

| Institution | City, Country (Romania) |
| --- | --- |
| Clinical Hospital ‘Dr Ion Cantacuzino’ Bucharest | Bucharest |
| Hospital MedLife Humanitas Cluj-Napoca | Cluj-Napoca |
| Medical office of Dr Saleh K. Majed | Craiova |
| Medical office of Dr Surpanelu Oana | Iasi |
| Natisan Medical Center | Pitesti |
| Gynecological office of Dr Rădulescu G. Mihaela Elena | Râmnicu Vâlcea |
| Medical office of Dr. Popescu | Sibiu |
| MediBlue Medical Clinic | Iasi |
| SC Pan Medical SRL | Sibiu |
| Medical clinic of Dr. Cioata Ionel Trifon | Timisoara |
| iMED Clinic | Sibiu |
| Tulcea County Emergency Hospital | Tulcea |
| Bradmed SRL | Targu Jiu |
| Gynecological office of Dr Ioana Trotea Targu Jiu | Targu Jiu |
| Medical office of Obstetrics and Gynecology of Dr Sirbu Daniela SRL | Timisoara |
| Dr Todorut Florina | Timisoara |
| Total sites, 16. |  |

Prospective data were collected from each patient, such as initial diagnosis, colposcopy examination results, re-epithelialization degree, vaginal pH value, vaginal symptoms, any worsening symptoms and adverse events.

### Ethical and regulatory aspects of the study

Written consent for study participation was collected from all patients. General Data Protection Regulation (GDPR) consent forms were collected from all patients. Due to legal considerations (GDPR directive effective from 21 May 2018 in all European Union countries), patients or their legal representatives have an absolute right to request that their data be removed from the study database. A Notified Body (ENTE CERTIFICATIONE MACCHINE SRL) reviewed the post marketing clinical follow-up plan, including ethical considerations. As this is a post-marketing clinical follow-up study, an Ethics Committee approval was not required, as per the regulations described below.

The study was conducted in accordance with the Guide to medical devices: ‘Post-market clinical follow-up studies’ (https://www.imdrf.org/sites/default/files/docs/imdrf/final/technical/imdrf-tech-210325-wng65.pdf) and the International Society for Pharmacoepidemiology (ISPE; 2015) Guidelines for ‘Good pharmacoepidemiology practices (GPP)’ (https://www.pharmacoepi.org/resources/policies/guidelines-08027/)

The collected data and study procedures were conducted in accordance with the ethical principles that have their origin in the Declaration of Helsinki.

The study followed the definition of the non-interventional (observational) study provided by the Guide to Good Pharmacovigilance Practices (GVP; 2017): Module VIII - Post-authorization safety studies (https://www.ema.europa.eu/en/documents/scientific-guideline/guideline-good-pharmacovigilance-practices-gvp-module-viii-post-authorisation-safety-studies-rev-3_en.pdf). The study followed the nature of the non-interventional (observational) studies mentioned in the ICH Harmonized Tripartite Guide Pharmacovigilance Planning E2E (ICH, 2004; https://database.ich.org/sites/default/files/E2E_Guideline.pdf).

Revision risk analysis was carried out in accordance with the medical device regulation, available from: https://www.anm.ro/en/dispozitive-medicale/regulamentele-europene-privind-dispozitivele-medicale. Data were stored according to Annex E of ISO 14155:2020 (https://www.iso.org/standard/). The study and its details are registered in www.clinicaltrials.gov under ID NCT05668806.

### Statistical consideration

All statistical analyses were performed using the Excel Analysis ToolPak, version 16.69.1, from Microsoft. P<0.05 was considered to indicate a statistically significant difference.

The quality and completeness of the collected data were preliminarily assessed in comparison with data analysis. No study participant was involved in any violation of inclusion/exclusion criteria. To examine the treatment significance over time, Fisher’s exact test was performed for categorical variables, and Mann-Whitney U test was employed to perform comparative analysis for variables non-normally distributed.

## Results

### Range of gynecological conditions and medical history of selected patients

The selected patients presented with various gynecological conditions consistent with the instructions for the use of the medical device. Some patients presented with other conditions with similar symptoms. As the evaluation of the performance of the device was a secondary objective, other conditions that would enable further investigations for expanding the use of the medical device were also selected (Table 2).

**Table 2.**
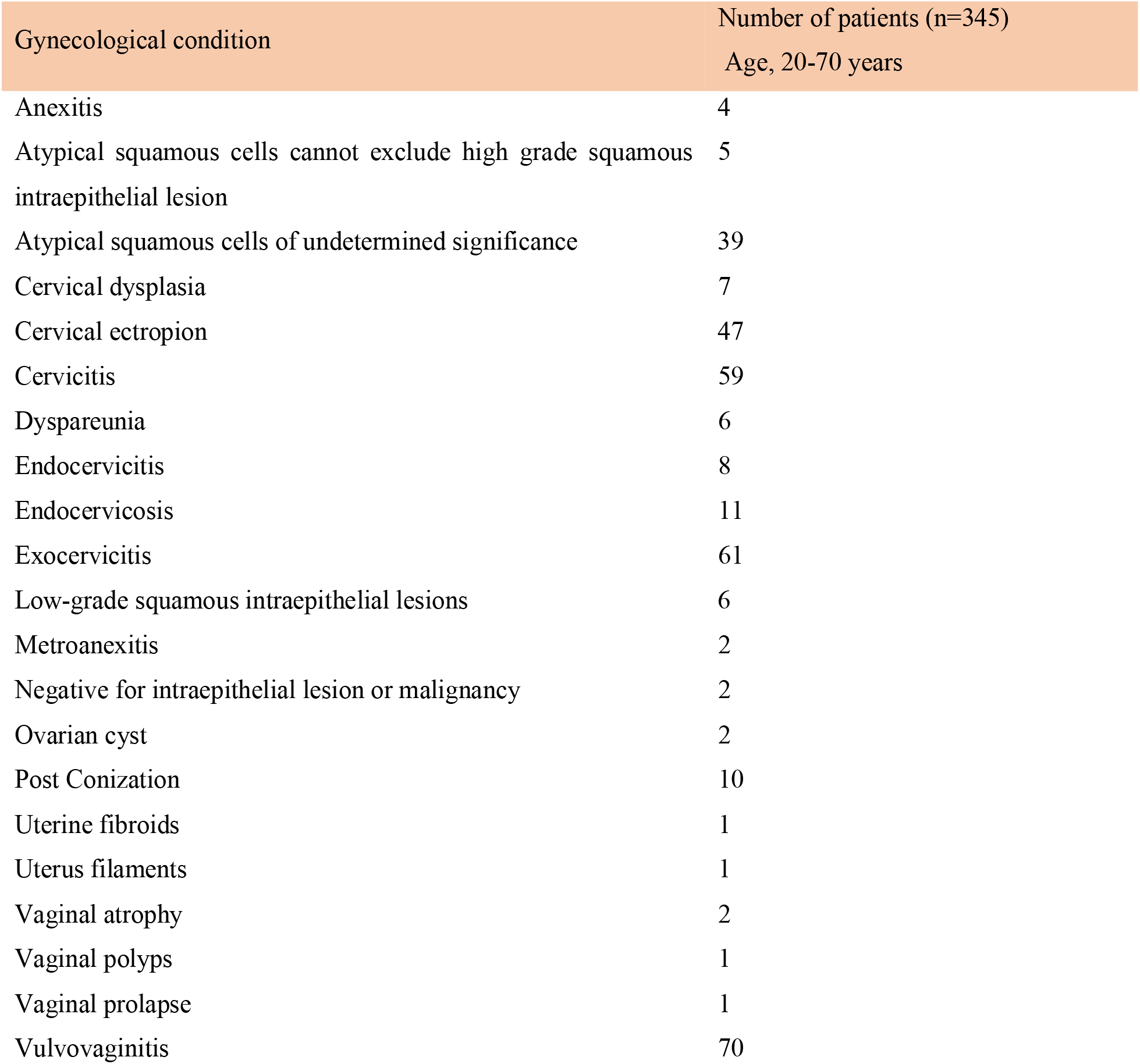
Medical history of selected patients

Subjects were followed undergoing treatment with Cerviron^®^ vaginal ovules for ∼3 months. Each enrolled subject visited the clinic four times, at 30-day intervals each. The study visit scheme is listed in figure 1. In the monotherapy group, 11 patients (5.23%) were considered dropouts due to the fact that they did not attend all the study visits and could not be evaluated. In the polytherapy group, 32 patients (23.7%) were considered dropouts (figure 1).

**Figure 1.**
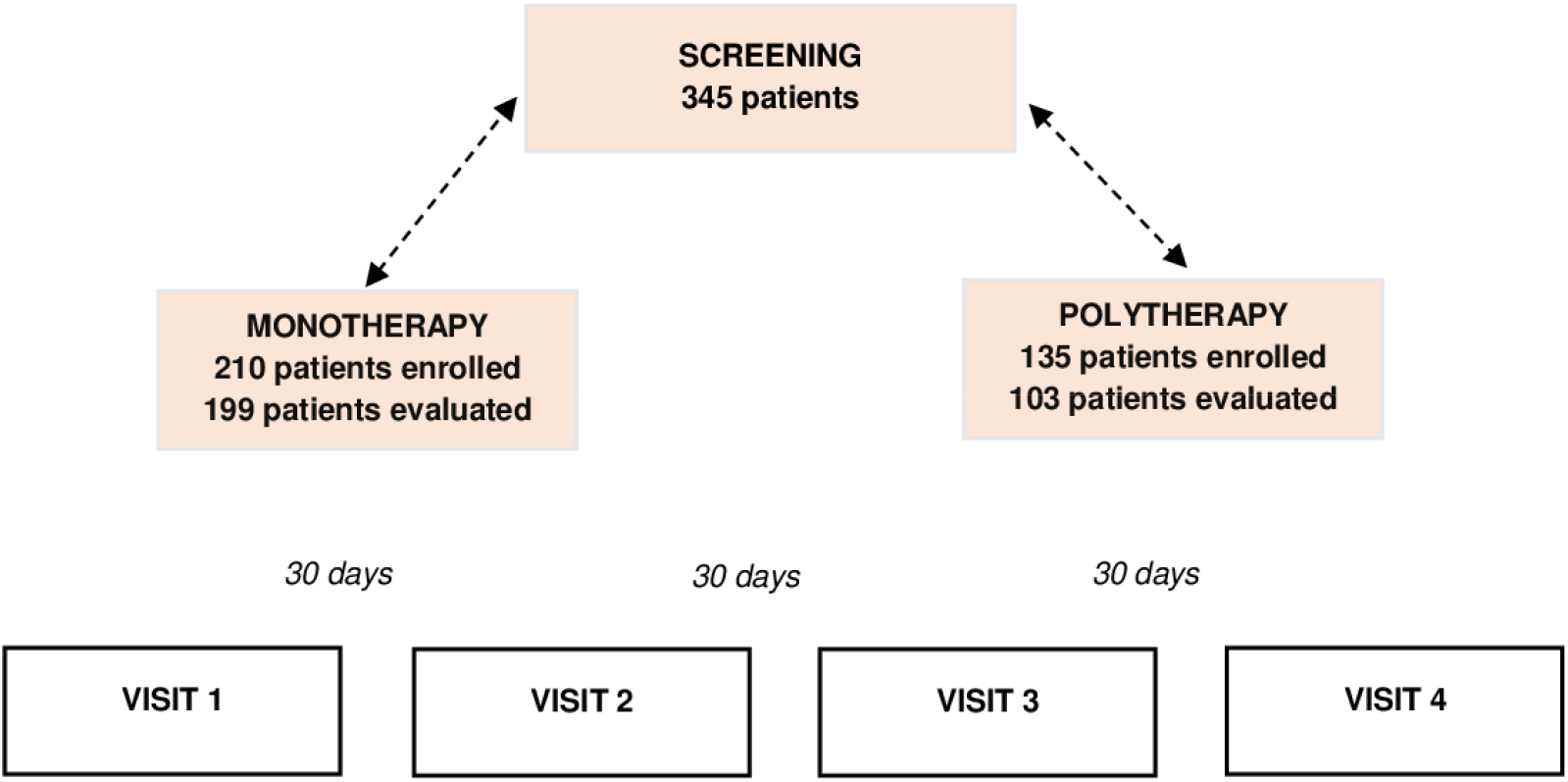
Schematic diagram for the present study.

### Primary objectives

The study primary objective was to assess the tolerability of the medical device by the number of possible adverse reactions observed during the treatment. In one patient treated exclusively with Cerviron^®^ ovules, during the routine gynecological examination one adverse reaction was noted, which was erythema. The erythema was followed by itching and abnormal vaginal discharge. The investigators determined that this was not related to treatment with the medical device. At the following visit, the reaction was absent.

Two other patients received combined treatment with antibiotics and Cerviron^®^ ovules. After 30 days, both patients reported an adverse reaction of itching and erythema. Investigators determined that the symptoms were consistent with the initial diagnosis. After 60 days, these reactions were absent in both patients. All the aforementioned incidents were linked to preexisting conditions and the investigators decided that they must not be regarded as adverse reactions to the medical device Cerviron^®^.

### Secondary objectives

#### Clinical performance assessed by the investigator by colposcopy

A significant proportion of 69.14% patients were rated with the indicator ‘normal aspect’ during colposcopy, as shown in figure 2.

**Figure 2.**
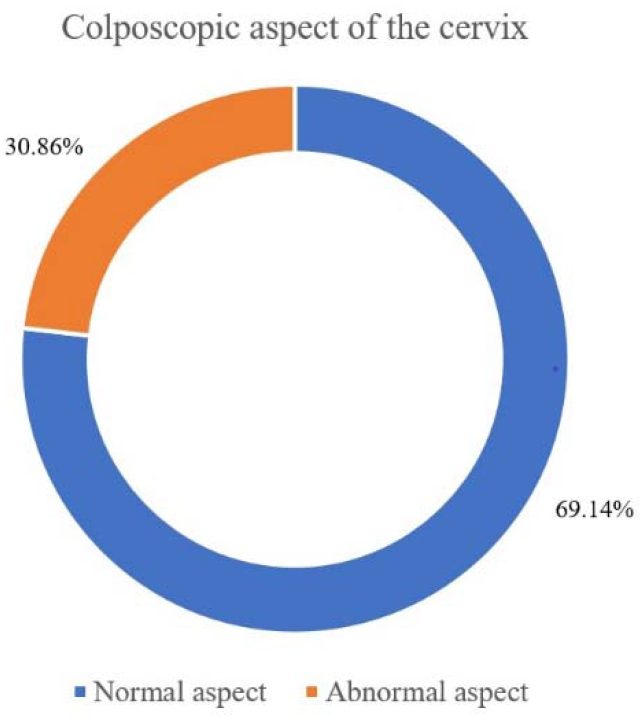
Clinical performance assessed by the investigator by colposcopy

#### Re-epithelialization degree of the cervical mucosa by gynecological examination

For only 302 patients out of 345, medical records evaluating the degree of epithelialization of the cervical mucosa were available. At the initial visit, 302 participants were assessed, of which 3.64% were rated with ‘complete epithelization’, 40.40% were rated with degree of ‘partial re-epithelialization’ and 55,96% were rated with ‘absent, ulcerations present’. At 30 days, 47.68% of the 302 women assessed, had a favorable degree of re-epithelialization, with only 4.30% rated as ‘absent, ulcerations present’. After 3 months, 73.17% of the 287 patients were rated with ‘complete epithelization’, as shown in table 3.

**Table 3.**
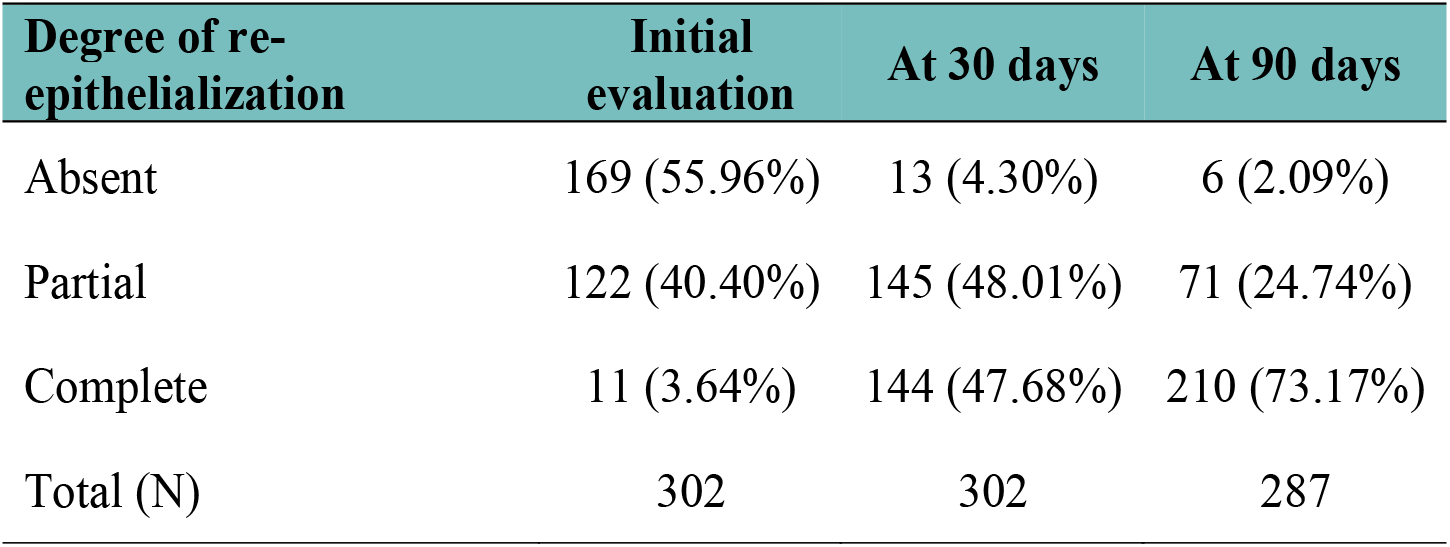
Degree of re-epithelization at initial evaluation, 30 and 90 days.

#### Vaginal pH level evaluation

It was observed that following a 3-month treatment with Cerviron®, 87.86% of the patients presented with a normal pH, as revealed in figure 3.

**Figure 3.**
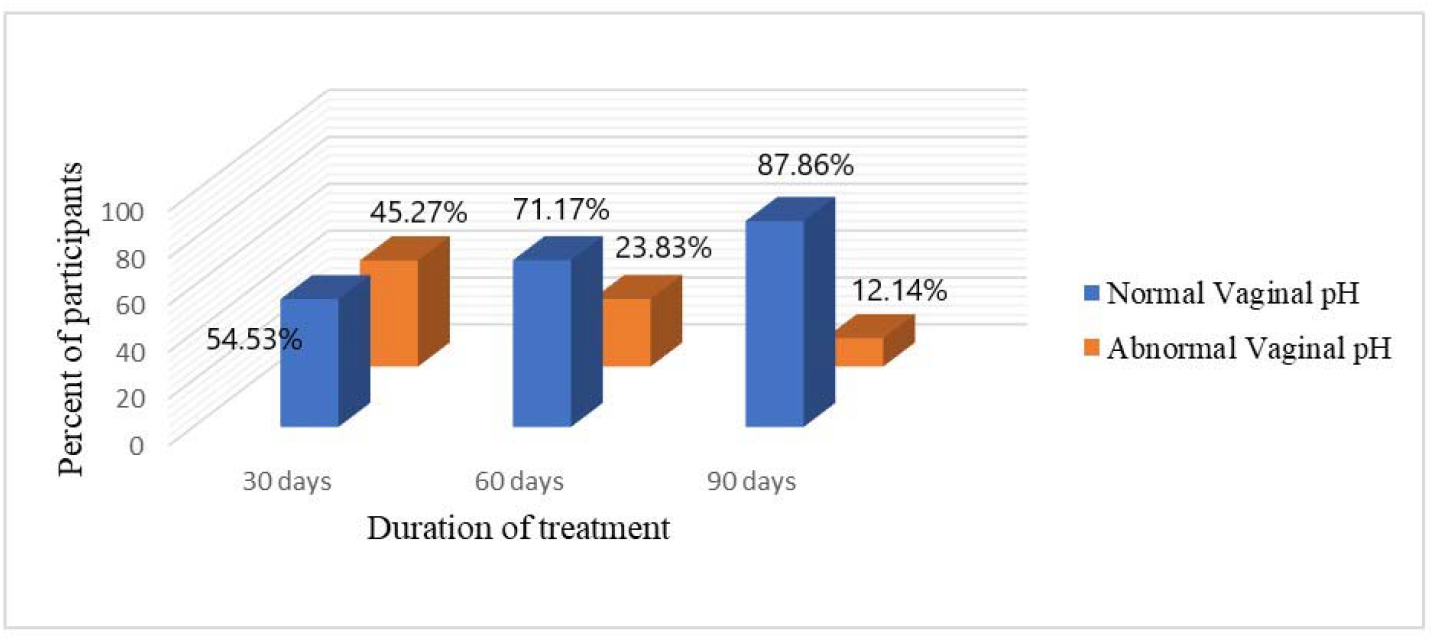
Changes in vaginal pH throughout the study (90 days).

#### Evaluation of the level of pain

Of the total number (n=302) of patients examined at visit 1, 46.67% reported mild pain levels and 28.00% had no pain at all. At the second visit, 68.49% of the population (207) rated pain severity as no pain or mild severity in 26.71%. A very high proportion (86.47%) was pain free at the last visit after 3 months of treatment with Cerviron^®^ (as shown in figure 4). Figure 4 also provides the corresponding data related to pain level across the treatment period.

**Figure 4.**
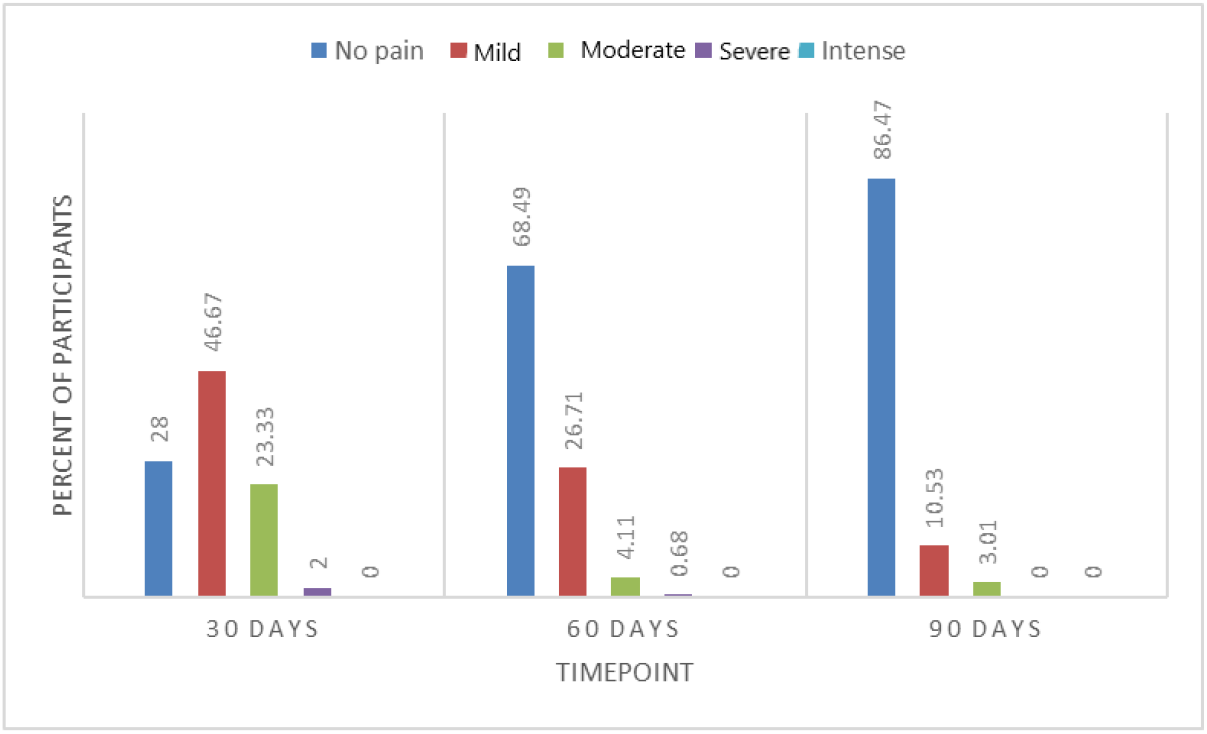
Evaluation of pain level between the study visits (90 days).

#### Evaluation of the level of vaginal bleeding

At the first visit, 302 women included in the study were evaluated, of whom 48.70% reported no bleeding observed after treatment with Cerviron® and 33.91% reported low amounts of vaginal bleeding. The efficacy of medical device was more clearly observed at visit 2, where 77.68% of patients experienced no bleeding at all and 1.79% experienced moderate or severe bleeding. After 3 months of treatment, this improvement wa noteworthy when 92.73% of patients reported no bleeding and only 7.27% of patients reported a mild level of bleeding, as observed in figure 5.

**Figure 5.**
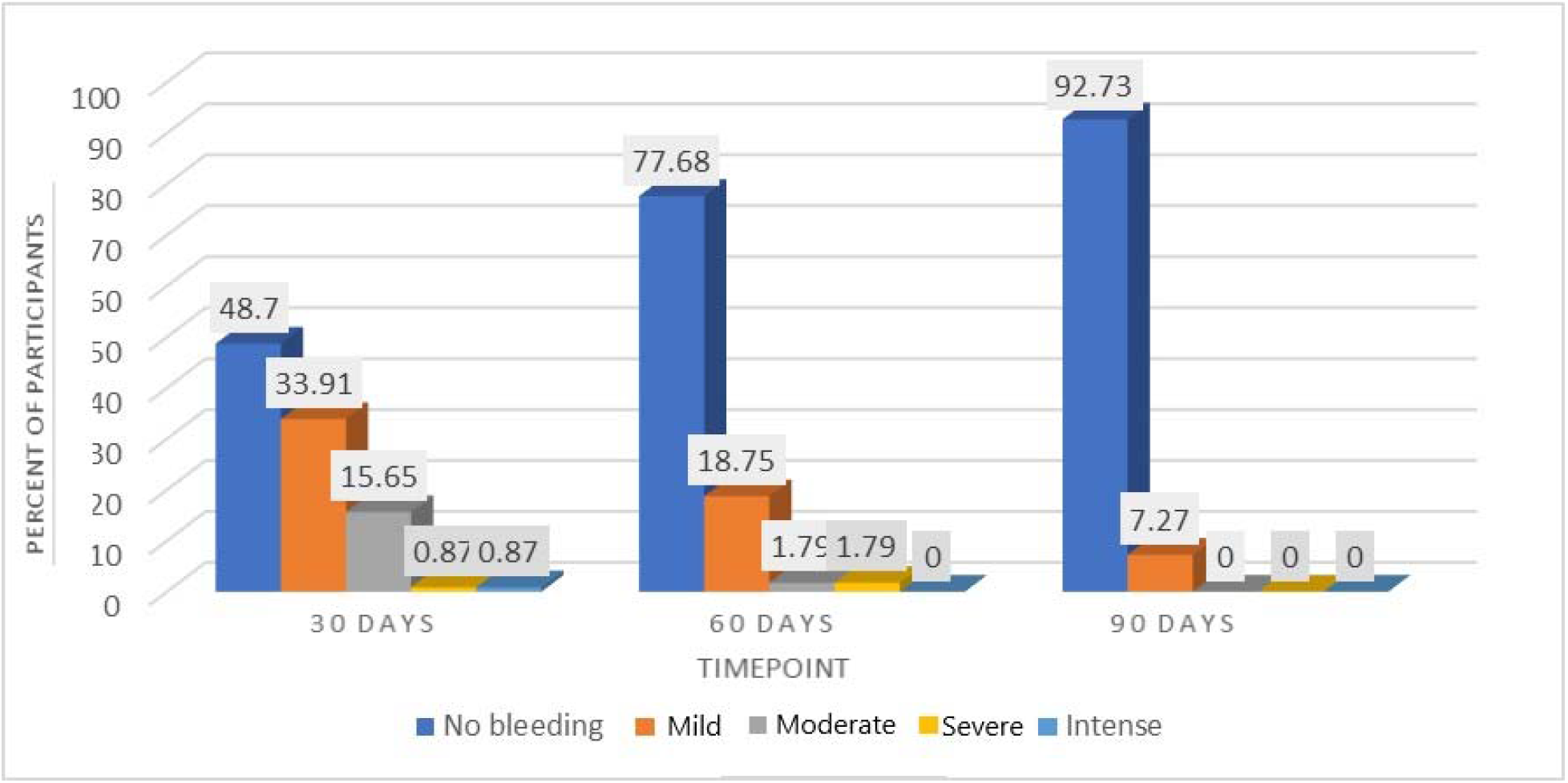
Evaluation of the level of vaginal bleeding throughout the study (90 days).

#### Degree of satisfaction after using Cerviron^®^ ovules

In terms of degree of satisfaction offered, participants in the largest population (72.25%) were reported to be ‘very satisfied’ with the medical device. The degree of satisfaction was measured by a 5-point Likert scale, as revealed in figure 6. (30).

**Figure 6.**
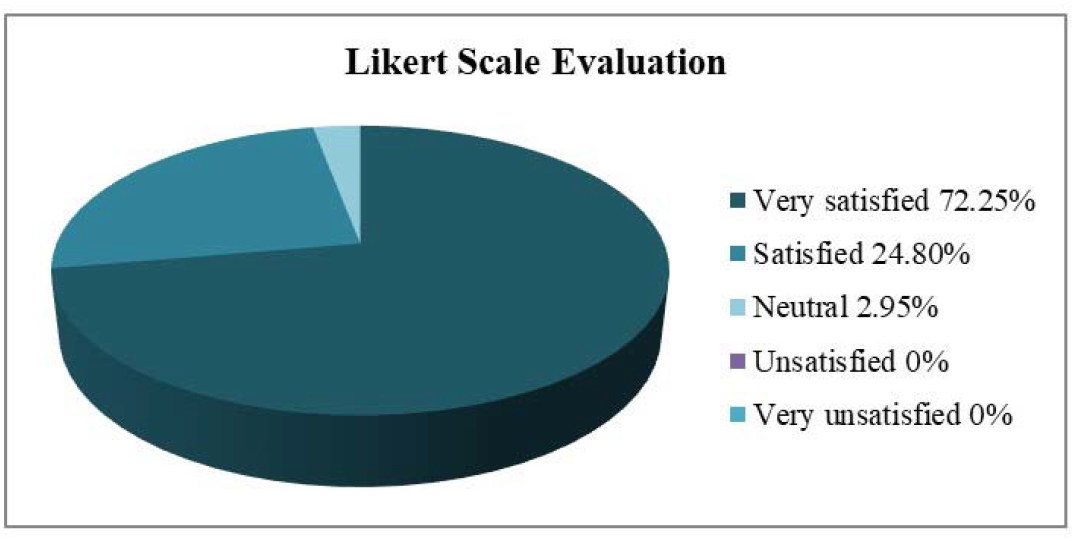
Likert Scale evaluation at the end of the study.

## Discussion

A very important aspect that influences the management of cervical ectopy is age. Cervical ectopy is predominant in young adolescents, with a prevalence of up to 80% in sexually active adolescents. (31) However, factors associated with the evolution and devolution of cervical ectopy are diverse and mechanisms have yet to be elucidated. Since ectopy is observed frequently in adolescent girls and pregnant women, it may vary in response to hormonal fluctuations observed in pregnancy and during contraception with combined oral contraceptives (32). Screening for C*hlamydia trachomatis* and *Neisseria gonorrhoeae* can prevent a range of gynecological conditions, such as urethritis, cervicitis, cervical erosions (33). Infection in the lower genital tract can result in upper genital tract complications, such as pelvic inflammatory diseases, ectopic pregnancy, chronic pelvic pain, and infertility in asymptomatic women, and transmission of infection during pregnancy and labor (34).

Cryotherapy (cryosurgery) of the cervix is an effective treatment for symptomatic cervical ectropion (35). Other surgical techniques have been applied for ablation of the squamocolumnar junction. Cauterization has been applied for this purpose since 1920, but it involves additional risks, such as burning lesions if the safeguard plate is not located correctly (36). Some specialists even consider the cervical ectropion as a normal finding that does not require treatment despite the red and inflamed appearance of the cervix. However, cervical ectropion is considered as one of the most common types of chronic cervicitis worldwide (37). Topical treatments for cervical ectropion, including cervical painting with gentian violet paint or microwave tissue coagulation, are still widely used (4). Although the debate is still ongoing, a harmonized treatment strategy would reduce complications of cervical ectropion, such as developing abnormal metaplasia and/or vaginal infection. Applying an intervention to treat cervical ectopy is recommended, especially when considering taking preventive action against cervical cancer. Ectopic cells are modified over time by squamous metaplasia and epithelialization, cervical infection with *Neisseria* gonorrhea or *Chlamidia trachomatis*, low pH values or trauma (38). Cerviron^®^ appears to have a very potent action in rebalancing the vaginal pH. In a previous clinical investigation, NCT04735705, vaginal pH values measured over 90 days showed that Cerviron^®^ restores altered vaginal pH. The difference in vaginal pH values between baseline versus 90 days, at a 5% significance level was statistically significant (P<0.05).

A literature review conducted by Machado *et al* indicated that treatment can be used to relieve occasional symptoms associated with ectopy, but does not support routine treatment for ectopy (12).

By contrast, a study conducted by Soares *et al*, showed a positive association between cervical ectopy and human papillomavirus, HIV, bacterial vaginosis, cervical epithelial atypia, postcoital bleeding, and desquamative inflammatory vaginitis (39). Recognition of cervical ectopy should alert the clinician to the possibility of a genital chlamydia infection. Opportunistic screening for chlamydia in young people should be offered to reduce the prevalence of infection and its sequelae. *Chlamydia trachomatis* is the most common bacterial sexually transmitted disease. In women, chlamydial infection usually presents as cervicitis, which can lead to up to 30 to 50% of pelvic inflammatory disease episodes. Pelvic inflammatory disease has significant reproductive sequelae, such as tubal infertility and ectopic pregnancy (40).

The main mechanism of action of the medical device is the dispersion of the substances in the vagina and the formation of a protective barrier that accelerates the natural healing process of the damaged epithelium. The effect of the medical device is obtained due to the presence of the Bismuth subgallate and vegetable collagen. Bismuth subgallate is an insoluble solution, with a very low bio-availability which will create a physical barrier over the affected area of the vaginal mucosa and therefore will not allow oxygen and pathogens to come in contact with it. In this way, the substances create the premises that allow the damaged tissue to heal naturally. The role of exogenous collagen is to be a ‘ sacrificial substrate’ in order to reduce excess metalloproteins that delay wound healing and to regulate the healing of the damaged tissue (14, 40-43). As such, Cerviron® supportive therapy may be prescribed in sexually active patients for at least 1-3 months, during10-15 consecutive days, to prevent these infections.

The findings in the present study revealed that a 90-day treatment (3 treatment sessions of 30 days each) with Cerviron vaginal ovules was beneficial in providing a complete degree of cervical epithelialization and reduced the multitude of the vaginal symptoms, including bleeding, inflammation, malodor, dysuria, dyspareunia, pain and leukorrhea. This result is consistent with previous clinical studies that included the same medical device, NCT04735705 and NCT04735718 (46).

In conclusion, the findings of the present study revealed that administration of Cerviron^®^ vaginal ovules provided a complete degree of epithelization in majority of patients attending a 90-days treatment. Moreover, subjects presenting a high grade of ulceration at the baseline visit, following treatment with Cerviron^®^ presented a complete degree of epithelization. Cerviron^®^ ovules are an exceptional adjuvant for acceleration of healing of cervical erosions. They favor the re-epithelialization of the damaged tissue and restoration of the initial colpo-ecosystem. Their topical application was observed to be effective in reducing unpleasant symptoms such as vaginal discharge, pelvic pain, and vaginal bleeding. In terms of adverse events, the medical device is considered safe. The only contraindication to Cerviron^®^ ovules has been determined as hypersensitivity to the active substance or to any of the excipients within the medical device.

## Data Availability

All data produced in the present study are available upon reasonable request to the authors.

## Acknowledgements

We would like to thank Mrs Florentina Liliana Calancea (MDX Research, Timisoara, Romania) for medical writing and Mr Alexandru Pinta (Labcorp Austria GmbH, Vienna Austria) for support in data management and statistical analysis.

## Availability of data and materials

The data that support the findings of this study are available from the corresponding author upon reasonable request.

## Authors’ contributions

IP, RP and ADT were involved in the study design, methodology and original draft preparation. IP and DTS performed the data collection and data analysis. RP, ADT and IP were involved in analysis and interpretation of data. EP and FD-P were involved in manuscript review and editing. IP, DTS and ADT participated in the interpretation of the study results. FD-P and EP confirm the authenticity of all the raw data. All authors approved the final version of the manuscript to be published.

## Ethics approval and consent to participate

Only participants that voluntarily provided their written consent to the collection of their study data were included. The collected data and study procedures were conducted in accordance with the ethical principles that have their origin in the Declaration of Helsinki. Ethics committee/IRB of Comisia Nationala de Bioetica a Medicamentului si a Dispozitivelor Medicale (CNBMDM) waived the ethical approval for this work, according to the European Medical Device Regulation MDR 2017/745, concerning data collected in the post-marketing clinical follow-up plan for medical devices, Annex XIV, part B.

## Patient consent for publication

Not applicable.

